# COVID-19-related disruptions to routine vaccination services in India: perspectives from pediatricians

**DOI:** 10.1101/2021.01.25.21250040

**Authors:** Anita Shet, Baldeep Dhaliwal, Preetika Banerjee, Kelly Carr, Andrea DeLuca, Carl Britto, Rajeev Seth, Bakul Parekh, GV Basavaraj, Piyush Gupta, Digant Shastri

## Abstract

**Background and Objective:** The COVID-19 pandemic has led to disruptions to routine immunization programs in India and around the world, setting the stage for potentially serious outbreaks of vaccine-preventable diseases.

**Methods:** We surveyed pediatric healthcare providers in India in 2 rounds in April-June and September 2020 to understand how COVID-19 control measures may have impacted routine vaccination.

**Results:** Respondents were predominantly pediatricians working in primary, secondary or tertiary healthcare centers, across 21 Indian states and two union territories. Among the 424 (survey 1) and 141 (survey 2) respondents, 33.4% and 7.8%, respectively, reported near complete suspension of vaccination services due to COVID-19. A 50% or greater drop in vaccination services was reported by 83.1% of respondents in June, followed by 32.6% four months later, indicating slow recovery of services. By September 2020, 83.6% were aware of updated guidelines on safe provision of immunization services, although awareness of specific catch-up vaccination plans was low, and 76.6% expressed concern about a vaccine coverage gap that could potentially lead to increased non-COVID-19 illnesses and deaths.

**Conclusions:** Pandemic-related disruptions to vaccination services were reported by pediatricians across India. Concerted efforts are needed from governing and academic groups to ensure that routine immunization and catch-up programs are implemented during this pandemic, which can sustain gains in vaccination coverage and provide a robust blueprint for the national roll-out of the COVID-19 vaccine.

## BACKGROUND

The COVID-19 pandemic has led to significant disruption in essential health services, including routine childhood immunizations[1, 2]. Evidence from previous epidemics have demonstrated that a temporary interruption of routine immunization services may lead to secondary health crises, such as outbreaks of vaccine-preventable diseases, amplifying morbidity and mortality[3]. Global interruptions in income, food supplies, and medical care can leave children at risk of becoming the unacknowledged victims of the current pandemic. The nationwide lockdown imposed by the Government of India on 24 March 2020 was formally lifted on 31 May 2020 with states continuing to monitor containment measures as needed. While the lockdown possibly slowed the spread of the SARS-CoV-2 virus and averted deaths due to COVID-19[4], it also resulted in concurrent crises in other aspects of health by taking a toll on healthcare provision and health-seeking activities[5]. Even services deemed as essential, including vaccination, were limited as a consequence of the lockdown and subsequent restrictions. India has made tremendous progress in reducing vaccination inequities in recent years[6]; however, these gains are at risk of being forfeited as a result of pandemic-related disruptions.

We surveyed pediatric healthcare providers in India at two timepoints to understand how COVID-19 control measures may have impacted routine vaccination at the peak of the lockdown and four months after the nationwide lockdown was lifted. The survey also explored barriers to healthcare provision and demand and identified innovative ways to regain losses in vaccination coverage.

## METHODS

The surveys were jointly developed by the Johns Hopkins Bloomberg School of Public Health (JHSPH) and the Indian Academy of Pediatrics (IAP). The first survey (survey 1) was launched virtually in April-June 2020 using the software platform Qualtrics, and was distributed to a network of pediatricians, primary care clinicians, and other care providers. The second survey (survey 2) was distributed in September 2020, after the nationwide lockdown had been lifted for three months. Distribution of both surveys and requests to complete them took place via email, text messages, telephone, in-person, and social media platforms such as WhatsApp. Survey topics included: perceptions of the impact of COVID-19 on caregiver vaccine-seeking behavior; assessment of routine vaccination services and vaccine campaigns; plans for resuming vaccination services; strategies employed to motivate vaccine uptake; and, perceived barriers caregivers and providers face in accessing and providing vaccinations. Surveys were anonymous and no personal identifiers were collected. This study was reviewed by the JHSPH Institutional Review Board (IRB) and approved as a non-human subjects research. Responses were collated in Qualtrics and analysed using Stata version 15.1.

## RESULTS

Survey 1 was circulated between 21 April and 11 June 2020, recording 424 responses representing 21 Indian states and two union territories. Majority of the respondents (96.0%, 407/424) were pediatricians who worked in predominantly urban and private primary, secondary or tertiary healthcare centers. Among these respondents, 86.6% (367/424) reported they were directly involved in providing vaccination services at their center.

Complete or partial suspension of immunization services at their respective centers was reported by 33.4% (142/424) of respondents (Table 1). Among vaccinations facilities, 83.1% (329/396) observed a >50% decrease in the number of children and families seeking vaccinations. Known interruption of vaccine campaigns and outreach programs for measles, rubella, and polio vaccinations were reported by 37.7% (160/424) of respondents. Only 38.7% (164/424) of respondents reported having a plan for catch-up vaccination once these restrictions were eased.

**Table 1:**
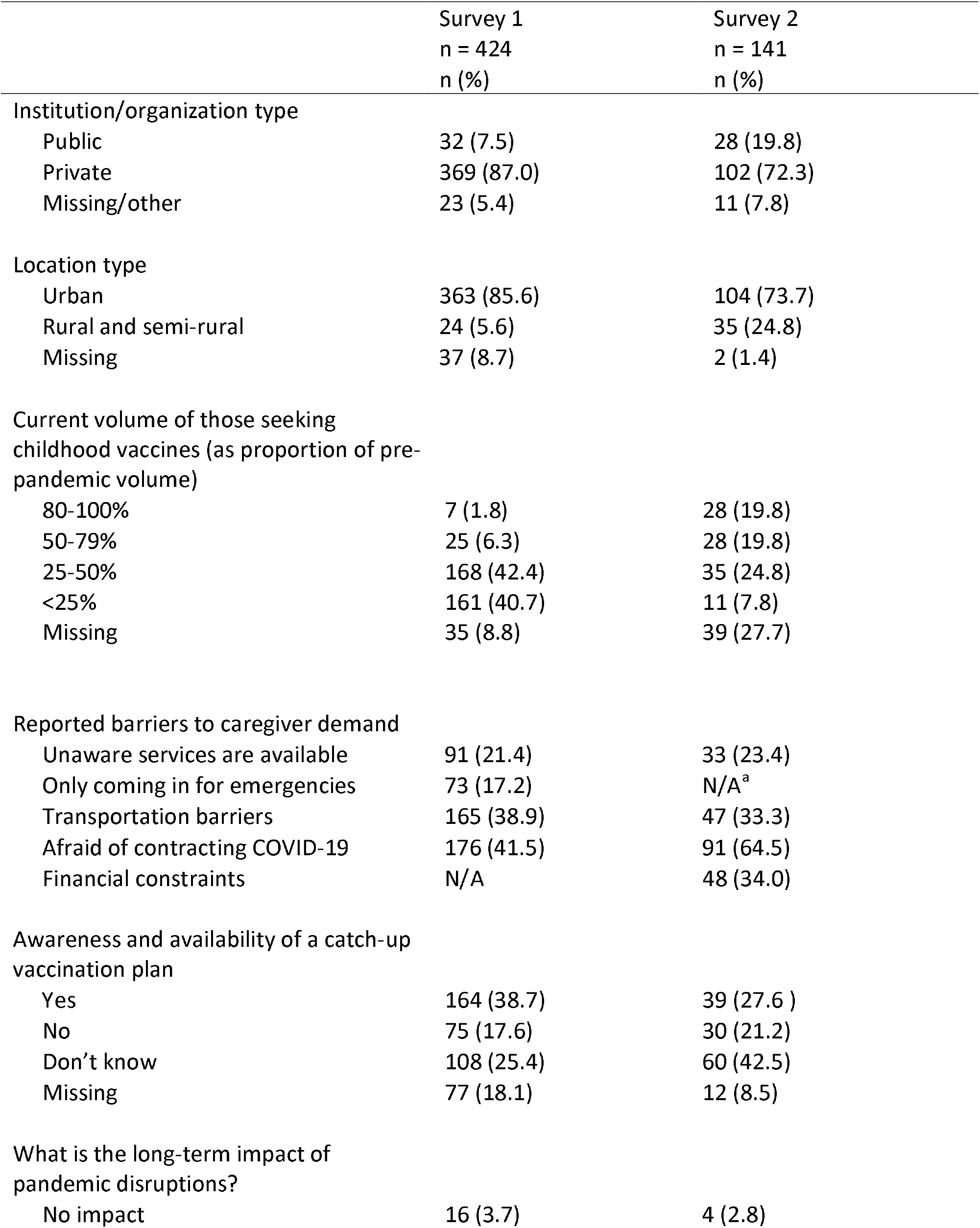
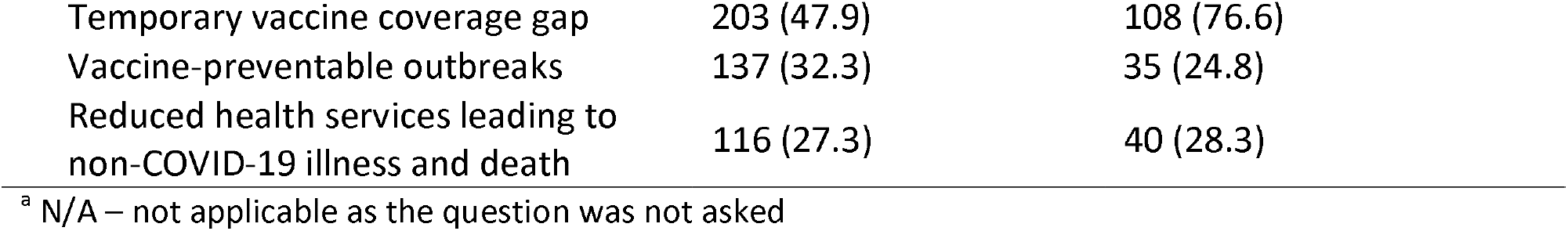
Survey respondents’ characterteristcs and perceptions around routine vaccination in the context of COVID-19, April-June 2020 (Survey 1) and September 2020 (Survey 2)

Survey 2 resulted in 141 responses between 8-25 September 2020. The characteristics of these respondents were similar to those from the first survey, with 92.9% (131/141) pediatricians, representing 21 states and one union territory. Respondents who did not participate in Survey 1 constituted 57.4% (81/141). Over half of respondents (51.7%, 73/141) reported that compared to pre-pandemic days, they were providing vaccinations less frequently. In Survey 2, only 32.6% (46/141) of respondents shared that they were seeing less than half the typical patient volume; this was an improvement from the high proportion of respondents who reported this drop in patient volume in Survey 1.

Pediatricians shared their perceptions of supply-side (healthcare availability) and demand-side barriers faced by caregivers. From survey 2, notable supply-side barriers were: low availability of healthcare workers; financial constraints; and, limited supplies, including PPE (Figure 1). Barriers to caregiver vaccine uptake included: low awareness of the availability of services; transportation limitations; fear of contracting COVID-19 from a clinical setting; and, financial contstraints (Table 1).

**Figure 1:**
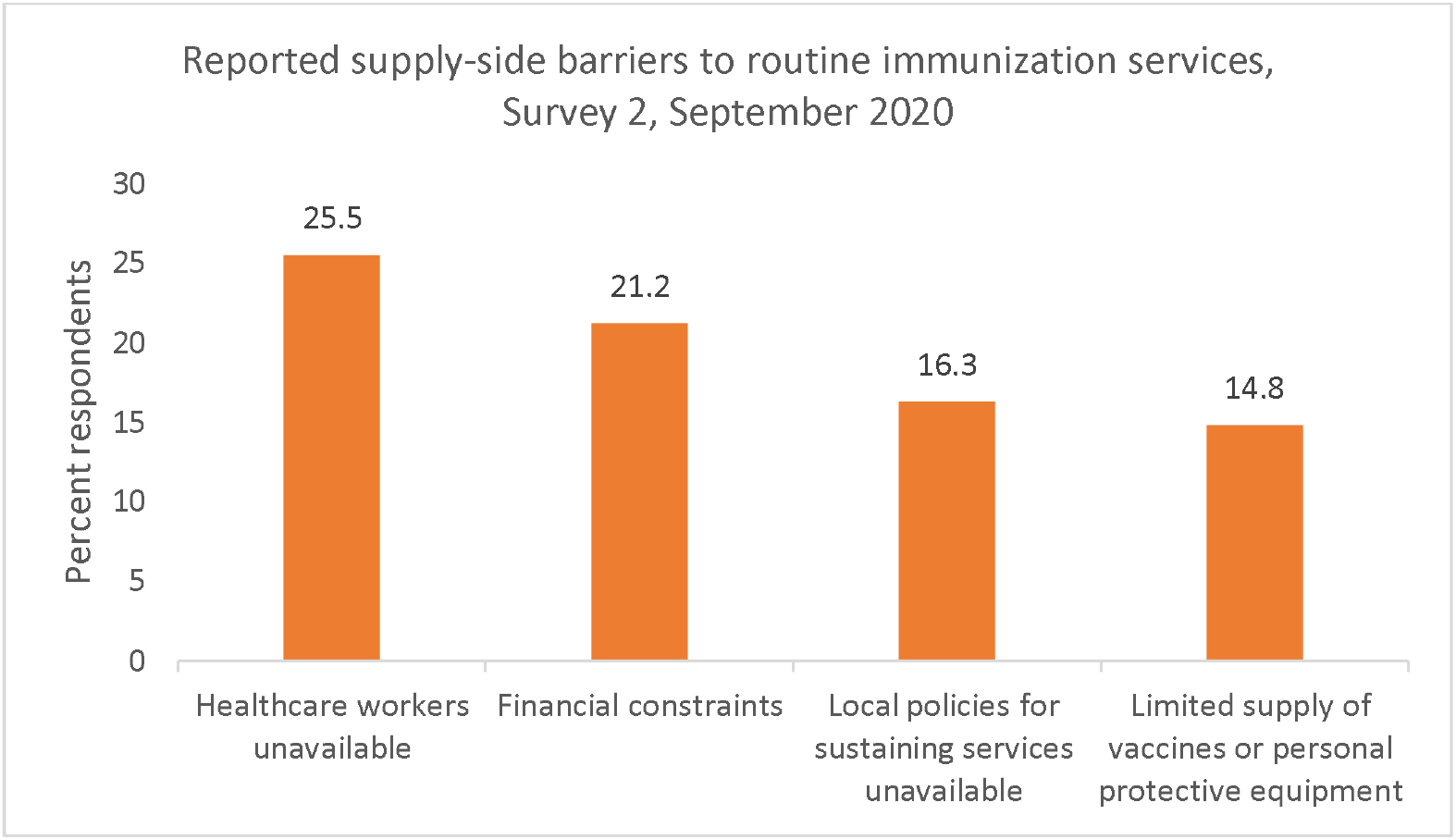
Reported supply-side barriers to routine immunization services, Survey 2, September 2020.

Among respondents, 61.7% (87/141) reported receiving guidelines on safe provision of vaccinations during the COVID-19 pandemic through the Indian Academy of Pediatrics (IAP). However, 42.5% (60/141) of respondents claimed they were not aware of state or district activities to prioritize vaccination catch-up visits. Reported suggestions from respondents for catch-up vaccinations included: setting up localized communication and catch-up campaigns; door-to-door outreach campaigns; use of text messaging immunization campaigns; and, implementing appointment-only, affordable immunization visits.

When asked about the long-term impact of these disruptions on child health, 47.9% (203/424) of respondents from Survey 1 and 76.6% (108/141) in Survey 2 believed that the lockdown and its after-effects would result in a vaccine coverage gap (Table 1). Over a quarter of respondents to both surveys reported that reduced health services may lead to non-COVID-19 illnesses and deaths; and a similar proportion believed that lockdown related restrictions would result in imminent outbreaks of vaccine-preventable diseases.

## DISCUSSION

Immunization and child health monitoring are essential services that should be prioritized to ensure sustainable benefits to society. Our survey findings from primary care providers and pediatricians in India indicate substantial vaccine service interruptions and concerns about the long-term impact of lockdowns. With approximately one-quarter of our respondents predicting increased morbidity and death as well as an increase in vaccine-preventable diseases, frontline clinicians are anticipating long-lasting effects of pandemic-related disruptions on their patient populations. The WHO reported major disruptions to vaccination services in countries around the world, and estimated that approximately 80 million children under the age of one were living in countries where routine immunization services was disrupted, and could potentially be at risk for developing a vaccine-preventable illness[2]. Responses from our survey respondents amplify a growing chorus around the globe calling for a focus on vaccine-preventable illnesses, even as COVID-19 cases grow worldwide. This comes at a pivotal time in India’s ongoing pursuit to improve immunization coverage. The national immunization program run by the government of India is one of the largest in the world, with an annual reach of over 26 million children and 29 million pregnant women[7]. Mission Indradhanush was launched in 2014 to further extend this reach and achieve full immunization for 90% of children. While remarkable progress has been made, there is also evidence of existing inequalities in coverage[8].

Early in the pandemic, soon after the lockdown was announced, there were major disruptions in health services, especially in women and children’s services. Movement restrictions were likely to have disrupted strategies utilized by Mission Indradhanush, including community mobilization, door-to-door campaigns, and monitoring events. The National Health Mission’s health management and information system reported a substantial decrease in routine immunization services relative to the previous year, indicating that in March 2020, at least 100,000 and 200,000 children missed their BCG and pentavalent (diphtheria, tetanus, pertussis, hepatitis B and Hemophilus influenzae type b) vaccines respectively[9, 10]. Researchers modelled different scenarios and used the Lives Saved Tool (LiST) to demonstrate that widespread disruption to health systems could lead to substantial increases in maternal and child deaths [11]. In India they estimated an additional 49,000 child deaths and 2,300 maternal deaths in a month attributable to severely disrupted services[11]. Applying the current population demographic data in India, estimates suggested that eventually over 27 million children will miss out on DTP vaccines and other health services, resulting in a 40% increase in child mortality over the next year[12]. Without targeted campaigns and effort, there is a legitimate risk for a reversal of gains made through national programs. If vaccination services are not restored and barriers to access are not addressed, disparities will become more pronounced and the number of zero-dose children will likely increase. India currently accounts for 2.1 million of the 20 million unvaccinated and under-vaccinated children globally (11%)[13], and the national lockdown has shown its potential to further exacerbate this problem.

Experiences from past outbreaks provide lessons on the indirect impacts which can be even more harmful to health. Analyses from the West African Ebola outbreak in 2014-2015 suggest that the increased number of deaths caused by other infections such as measles, HIV and tuberculosis attributable to health system failures exceeded deaths from Ebola[3, 14]. A sustained period of disrupted immunization can result in the accumulation of susceptible individuals which in turn can lead to disease outbreaks[15]. Given the disruptions and the realization of the dire consequences, the Government of India declared immunization an essential health service in April 2020, and issued guidelines for states to resume routine immunization services[16]. In June 2020 India began a phased reopening of the economy, and the resumption of immunization activities were appropriately structured based on local COVID-19 infection rates and restrictions. These activities were based on the WHO guidance urging nations to continue providing essential services along with COVID-19 mitigation and treatment measures in order to maintain public trust and minimize morbidity and mortality[17].

Coordinated campaigns across India targeting children who missed critical routine vaccinations during the national lockdown, as well as targeting low-coverage areas, could prevent additional public health disasters. Prioriting measles vaccine catch up would be most prudent given the outbreak potential with even marginal reduction in heard immunity[18]. Planning catch-up campaigns now is essential, so providers can minimize the time children are at risk for vaccine-preventable diseases. Vaccination catch-up sessions could institute innovative strategies such as implementing appointment-only visits, minimizing overcrowding, separating immunization visits from sick children visits, prioritizing robust communication efforts which address caregivers’ fears of contracting COVID-19, and reminders to caregivers of the importance of routine vaccinations[19].

The Government of India has recently approved COVID-19 vaccines, and the nation has embarked on one of the largest and most ambitious immunization campaigns in the world. Although children will not receive the COVID-19 vaccine at this time, their caregivers and healthcare providers who will receive the vaccine, should be provided with targeted messages and reminders for childhood routine immunizations. In addition, liaising routine immunization campaigns with the COVID-19 vaccine rollout, particularly in hard-to-reach areas would be beneficial, given India’s vast human resources and immunization experience.

There are several limitations in this study. Immunization is predominantly offered by the public health sector in India, while the private health sector typically covers <10% of the population[20, 21]. Our results reflect a small proportion of the health care system in India, and may not represent effects in government and rural areas. Nonetheless, the private sector represents a substantial number of children in India, and can play an important role in supporting immunization activities undertaken by the government as seen in studies around the world[22]. The sample size was relatively small, and there may be selection bias as only those pediatricians interested in immunization may have responded. There were slight differences in the two rounds of the survey, as questions were modified for the second round based on new learnings, and the number of responses were lower in the second round.

There is growing evidence that the risk-benefit ratio is decidedly in favor of continuing vaccination services even when considering consequences of doing so during the pandemic[23]. Gaining provider insights on effective strategies is essential to establishing context-specific mechanisms to prioritize catch-up for missed vaccines, as well as ensure there is infrastructure in place for roll-out of the recently approved COVID-19 vaccine. The way forward should include an increased focus on catch-up campaigns, strong government engagement, effective surveillance, modelling analyses based on varying scenarios, and clear public health messaging, as these are critical next steps to ensure restoration of immunization and essential services for women and children.

## Data Availability

The authors confirm that the data supporting the findings of this study are available within the article [and/or] its supplementary materials. Derived data supporting the findings of this study are available from the corresponding author [AS] on request.

## REFERENCES

1. Chandir S, Siddiqi DA, Setayesh H, Khan AJ. Impact of COVID-19 lockdown on routine immunisation in Karachi, Pakistan. The Lancet Global health. 2020;8(9):e1118–e20.

2. World Health Organization, Special feature: immunization and COVID-19. June 2020. Available at: https://www.who.int/immunization/monitoring_surveillance/immunization-and-covid-19/en/.

3. Takahashi S, Metcalf CJ, Ferrari MJ, Moss WJ, Truelove SA, Tatem AJ, et al. Reduced vaccination and the risk of measles and other childhood infections post-Ebola. Science (New York, NY). 2015;347(6227):1240–2.

4. Laxminarayan R, Jameel S, Sarkar S. India’s Battle against COVID-19: Progress and Challenges. The American journal of tropical medicine and hygiene. 2020;103(4):1343–7.

5. The L. COVID-19 in India: the dangers of false optimism. Lancet. 2020;396(10255):867.

6. Gurnani V, Haldar P, Aggarwal MK, Das MK, Chauhan A, Murray J, et al. Improving vaccination coverage in India: lessons from Intensified Mission Indradhanush, a cross-sectoral systems strengthening strategy. BMJ (Clinical research ed). 2018;363:k4782.

7. National Health Mission: Immunization. Ministry of Health and Family Welfare, Government of India. Available at: https://nhm.gov.in/index1.php?lang=1&level=2&sublinkid=824&lid=220.

8. Intensified Mission Indradhanush 2.0: Coverage report. 25 September 2020. Available at: https://imi2.nhp.gov.in/report/coverage Accessed 20 November 2020.

9. National Health Mission Health Management Information System, Ministry of Health and Family Welfare, Government of India. Available at: https://nrhm-mis.nic.in/SitePages/Home.aspx. Accessed on 15 November 2020.

10. Rukmini S. How covid-19 response disrupted health services in rural India. Mint. 2020. https://www.livemint.com/news/india/how-covid-19-response-disrupted-health-services-in-rural-india-11587713155817.html. Accessed on 15 November 2020.

11. Roberton T, Carter ED, Chou VB, Stegmuller AR, Jackson BD, Tam Y, et al. Early estimates of the indirect effects of the COVID-19 pandemic on maternal and child mortality in low-income and middle-income countries: a modelling study. The Lancet Global health. 2020;8(7):e901–e8.

12. Global Financing Facility. Preserve Essential Health Services During the COVID-19 Pandemic: India. 2020. https://idronline.org/user-content/uploads/2020/06/India-Covid-Brief_GFF.pdf. Accessed 18 November 2020.

13. Progress and challenges with achieving universal immunization coverage. 2019 WHO/UNICEF Estimates of national immunization coverage. 15 July 2020. Available at: https://www.who.int/immunization/monitoring_surveillance/who-immuniz.pdf Accessed on 10 Novemebr 2020.

14. Elston JW, Cartwright C, Ndumbi P, Wright J. The health impact of the 2014-15 Ebola outbreak. Public health. 2017;143:60–70.

15. Truelove SA, Graham M, Moss WJ, Metcalf CJE, Ferrari MJ, Lessler J. Characterizing the impact of spatial clustering of susceptibility for measles elimination. Vaccine. 2019;37(5):732–41.

16. Immunization services during and post COVID-19 outbreak. Ministry of Health and Family Welfare, Government of India, 15 April 2020. Available at: https://www.mohfw.gov.in/pdf/3ImmunizationServicesduringCOVIDOutbreakSummary150520202.pdf.

17. World Health Organization. Maintaining essential health services: operational guidance for the COVID-19 context interim guidance, 1 June 2020. Available at: https://www.who.int/publications/i/item/covid-19-operational-guidance-for-maintaining-essential-health-services-during-an-outbreak Accessed 18 November 2020.

18. Masters NB, Eisenberg MC, Delamater PL, Kay M, Boulton ML, Zelner J. Fine-scale spatial clustering of measles nonvaccination that increases outbreak potential is obscured by aggregated reporting data. Proceedings of the National Academy of Sciences of the United States of America. 2020;117(45):28506–14.

19. Oyo-Ita A, Wiysonge CS, Oringanje C, Nwachukwu CE, Oduwole O, Meremikwu MM. Interventions for improving coverage of childhood immunisation in low- and middle-income countries. The Cochrane database of systematic reviews. 2016;7(7):Cd008145.

20. Sharma A, Kaplan WA, Chokshi M, Zodpey SP. Role of the private sector in vaccination service delivery in India: evidence from private-sector vaccine sales data, 2009-12. Health policy and planning. 2016;31(7):884–96.

21. Sarveswaran G, Krishnamoorthy Y, Sakthivel M, Vijayakumar K, Priyan S, Thekkur P, et al. Preference for Private Sector for Vaccination of Under-Five Children in India and Its Associated Factors: Findings from a Nationally Representative Sample. Journal of tropical pediatrics. 2019;65(5):427–38.

22. Levin A, Kaddar M. Role of the private sector in the provision of immunization services in low- and middle-income countries. Health policy and planning. 2011;26 Suppl 1:i4–12.

23. Abbas K, Procter SR, van Zandvoort K, Clark A, Funk S, Mengistu T, et al. Routine childhood immunisation during the COVID-19 pandemic in Africa: a benefit-risk analysis of health benefits versus excess risk of SARS-CoV-2 infection. The Lancet Global health. 2020;8(10):e1264–e72.

