## Supplementary material for "COVID-19-related disruptions to routine vaccination services in India: perspectives from pediatricians": Table 1

*Table 1: Survey respondents’ characteristics and perceptions around routine vaccination in the context of COVID-19, April-June 2020 (Survey 1) and September 2020 (Survey 2)*

|  | Survey 1  n = 424 | | | Survey 2  n = 141 |
| --- | --- | --- | --- | --- |
|  | n (%) | | | n (%) |
| Institution/organization type |  | | |  |
| Public | 32 (7.5) | | | 28 (19.8) |
| Private | 369 (87.0) | | | 102 (72.3) |
| Missing/other | 23 (5.4) | | | 11 (7.8) |
| Location type |  | | |  |
| Urban | 363 (85.6) | | | 104 (73.7) |
| Rural and semi-rural | 24 (5.6) | | | 35 (24.8) |
| Missing | 37 (8.7) | | | 2 (1.4) |
| Current volume of those seeking childhood vaccines (as proportion of pre-pandemic volume) | |  |  | |
| 80-100% | | 7 (1.8)^b^ | 28 (19.8) | |
| 50-79% | | 25 (6.3)^b^ | 28 (19.8) | |
| 25-50% | | 168 (42.4) ^b^ | 35 (24.8) | |
| <25%  Missing | | 161 (40.7) ^b^  35 (8.8) ^b^ | 11 (7.8)  39 (27.7) | |
| Reported barriers to caregiver demand | |  |  | |
| Unaware services are available | | 91 (21.4) | 33 (23.4) | |
| Only coming in for emergencies | | 73 (17.2) | N/A^a^ | |
| Transportation barriers | | 165 (38.9) | 47 (33.3) | |
| Afraid of contracting COVID-19 | | 176 (41.5) | 91 (64.5) | |
| Financial constraints | | N/A | 48 (34.0) | |
| Awareness and availability of a catch-up vaccination plan | |  |  | |
| Yes | | 164 (38.7) | 39 (27.6 ) | |
| No | | 75 (17.6) | 30 (21.2) | |
| Don’t know | | 108 (25.4) | 60 (42.5) | |
| Missing | | 77 (18.1) | 12 (8.5) | |
| What is the long-term impact of pandemic disruptions? | |  |  | |
| No impact | | 16 (3.7) | 4 (2.8) | |
| Temporary vaccine coverage gap | | 203 (47.9) | 108 (76.6) | |
| Vaccine-preventable outbreaks | | 137 (32.3) | 35 (24.8) | |
| Reduced health services leading to non-COVID-19 illness and death | | 116 (27.3) | 40 (28.3) | |

^a^ N/A – not applicable as the question was not asked

^b^ Denominator of 396 as question was asked to a subset of respondents
