## Supplementary material for "COVID-19-related disruptions to routine vaccination services in India: perspectives from pediatricians": Figure 1

*Figure 1: Reported supply-side barriers to routine immunization services, Survey 2, September 2020*

Reported supply-side barriers to routine immunization services, Survey 2, September 2020

|  | Percent respondents |
| --- | --- |
| Healthcare workers unavailable | 25.5 |
| Financial constraints | 21.2 |
| Local policies for sustaining services unavailable | 16.3 |
| Limited supply of vaccines or personal protective equipment | 14.8 |
